# Non-obstetric risk factors for deformational plagiocephaly: A systematic review and meta-analysis

**DOI:** 10.1101/2023.10.01.23296397

**Authors:** Dr Christopher Hillyar, Natalie Bishop, Frances J Bell-Davies, Juling Ong

**Author notes:** **Corresponding author** Dr Christopher Hillyar.

## Abstract

Plagiocephaly is defined as asymmetrical distortion of the skull resulting in an oblique trapezoid or parallelogram head shape. In the absence of skull growth restriction due to craniosynostosis, deformational plagiocephaly (DP) is caused by deformational forces acting on one side of the back of the head which distorts the normal symmetry of the skull. The aims of this systematic review and meta-analysis were to critically assess the evidence for non-obstetric risk factors for DP and to make evidence-based recommendations for reducing the prevalence of DP. A search of PubMed and Web of Science was performed covering 21 August 2010 to 21 August 2022. The searches yielded 159 articles, of which 18 articles were eligible for inclusion in this study. 43 non-obstetric factors were identified. Of these, a total of 17 factors were associated with DP. With the notable exceptions of maternal age, mechanical ventilation and tummy time, these associations were either supported by non-conflicting evidence or a meta-analysis that resolved conflicting evidence into a significant association. Thirteen factors had significant odds ratios that ranged from 1.10 (mechanical ventilation) to 7.15 (insufficient vitamin D intake). Of the five factors assessed by meta-analysis (male gender, reaching fewer motor milestones by six months of age, maternal education level, head position preference, sleeping position), only one (male gender) was associated with significant inter-study heterogeneity. No evidence of publication bias was detected. In summary, this study provides the most comprehensive meta-analytic assessment of non-obstetric factors associated with DP published to date. It provides 13 evidence-based recommendations which can be adopted by healthcare systems globally, to reduce the prevalence of DP and its impact on child development.

## Introduction

Plagiocephaly is defined as asymmetrical distortion of the skull resulting in an oblique trapezoid or parallelogram head shape when viewed from the vertex position in the axial plane [1]. The severity of skull asymmetry can range from minimal focal flattening on one side of the cranial vault to severe deformation affecting the entire cranial vault, skull base and facial skeleton. Plagiocephaly arises via two main mechanisms: premature fusion of one or more of the cranial sutures (craniosynostosis) or external mechanical forces acting on the cranial vault which result in a distortion of the normally symmetric craniofacial skeleton (deformational plagiocephaly (DP)).

In craniosynostotic plagiocephaly involving any of the paired coronal or lambdoid sutures, restriction of skull vault growth occurs perpendicular to the fused suture (Virchows Law) [2, 3]. Isolated craniosynostosis involving premature fusion of a single coronal suture results in an anterior plagiocephaly with brow retrusion on the affected side, and similarly, craniosynostosis of a single lambdoid suture will restrict posterior cranial growth on the same side. The asymmetry is often accentuated as the remaining unfused sutures expand to enable accommodation of the rapidly growing infant brain. The majority of patients with craniosynostosis do not have an identifiable genetic cause but this proportion is increased in patients with more than one suture involved [2, 3].

Alternatively - and far more commonly - plagiocephaly is caused by deformational forces acting on one side of the back of the head which distorts the normal symmetry of the skull in the absence of skull growth restriction due to craniosynostosis [4]. This deformity is characterised by an anterior asymmetric deformation of craniofacial skeleton in the parasagittal plane often described as a parallelogram type deformity. This presents clinically as an ipsilateral occipital flattening, anterior displacement of the ear, temperomandibular joint and frontal bossing [2–5]. The external forces applied to the head that result in flattening of the posterior neurocranium have given rise to the synonym “flat head syndrome” [6]. The importance of external forces can be seen in the close relationship between DP and sleeping position [7], among other factors that may influence external head forces [8–12].

Since the 1980s, ‘Back to Sleep’ (BTS) campaigns have successfully publicised the benefits of supine sleeping for reducing the risk of sleep-related death, including sudden infant death syndrome (SIDS) [13]. Although BTS reduced the incidence of SIDS by 40%, an undesirable consequence has been an increase in the prevalence of skull deformities, including a 600% increase in the incidence of DP [14, 15]. The majority of cases of DP will resolve without intervention and surgical treatment is not required [16]. However, in a subset of children DP persists beyond teenage years (prevalence of DP: 1.1% of 12 to 17 year olds), increasing the risk for developmental and psychological issues [17–19]. Although physiotherapy and helmet therapy may play a role in improving head shape and limiting other long-term effects [20, 21], understanding the factors that increase the risk of DP may help to prevent DP from developing. The aims of this systematic review and meta-analysis were to critically assess the evidence for risk factors for DP and to make evidence-based recommendations for reducing the prevalence of DP. This article was previously presented as a meeting abstract at the 2021 Royal College of Paediatrics and Child Health Conference on June 15, 2021

## Materials & Methods

The study protocol, analysis and reporting was conducted in-line with the Preferred Reporting Items for Systematic Reviews and Meta-Analyses (PRISMA) 2020 guidelines [22–24]. The protocol was registered with PROSPERO (Registration number: CRD42020204979), accessible online: https://www.crd.york.ac.uk/prospero/display_record.php?RecordID=234979.

### Search strategy

A search of PubMed and Web of Science was performed covering 21 August 2010 to 21 August 2022 (this included an initial search from 21 August 2010 to 20 August 2020 and an update search from 21 August 2010 to 20 August 2022 to ensure the list of included studies was as up-to-date as possible). The combination of PubMed and Web of Science provide >97.5% coverage of published literature [25, 26]. Additional databases, hand searching and grey literature were not included to balance comprehensive coverage with a pragmatic approach to ensure the study was completed with limited available resources. Search terms included “plagiocephaly” AND “risk factor”.

### Study eligibility

Study abstracts were screened and assessed for relevance. Specifically, original studies were included if they assessed risk factors for DP. Non-English language, non-human subjects, and low-quality or non-original studies (meeting abstracts, reviews, case series, case reports, and editorials) were excluded. Full-text review was performed after screening. Studies not reporting non-obstetric risk factors for DP were excluded. Preventative measures such as tummy time were included. However, treatments such as physical therapy and helmet therapy were not included.

### Data extraction and reporting

Data extraction and reporting followed PRISMA guidelines. Data from eligible full-text articles were extracted by three investigators (CRH, NB, AN). The main outcomes of interest were odd ratios (ORs) and risk ratios (RRs) (with 95% confidence intervals (CIs)), or significant associations, for risk factors for DP (or biomarkers of DP, e.g. oblique diameter difference index (ODDI)).

### Meta-analysis

Meta-analysis of OR/RR for factors associated with DP was performed. The inconsistency index (Ι2) and a Q-statistic for chi-squared significance for specific degrees of freedom (df) were calculated to assess inter-study heterogeneity. Both fixed effects and random effects were reported. P-values for 95% CIs were calculated. Microsoft Excel and GraphPad Prism were used for statistical analysis.

### Funnel plots

Publication bias was assessed using funnel plots of OR/RR for DP risk factors against study precision (1/standard error (SE)). Linear regression was utilised to assess Egger’s asymmetry, with a P-value of <0.05 indicating publication bias [27].

## Results

The searches yielded 159 articles combined from PubMed and Web of Science. After removing 36 duplicates, 124 articles were screened based on abstract content. Of these 124 articles, 77 were not relevant. Full-text screening of the remaining 47 articles resulted in exclusion of 20 irrelevant articles (did not report non-obstetric risk factors for deformational plagiocephaly), three non-English articles, and five articles which were not accessible. This resulted in 19 articles eligible for inclusion in this study (Figure *1*). The characteristics of eligible studies are illustrated in Table *1*.

**Figure 1:**
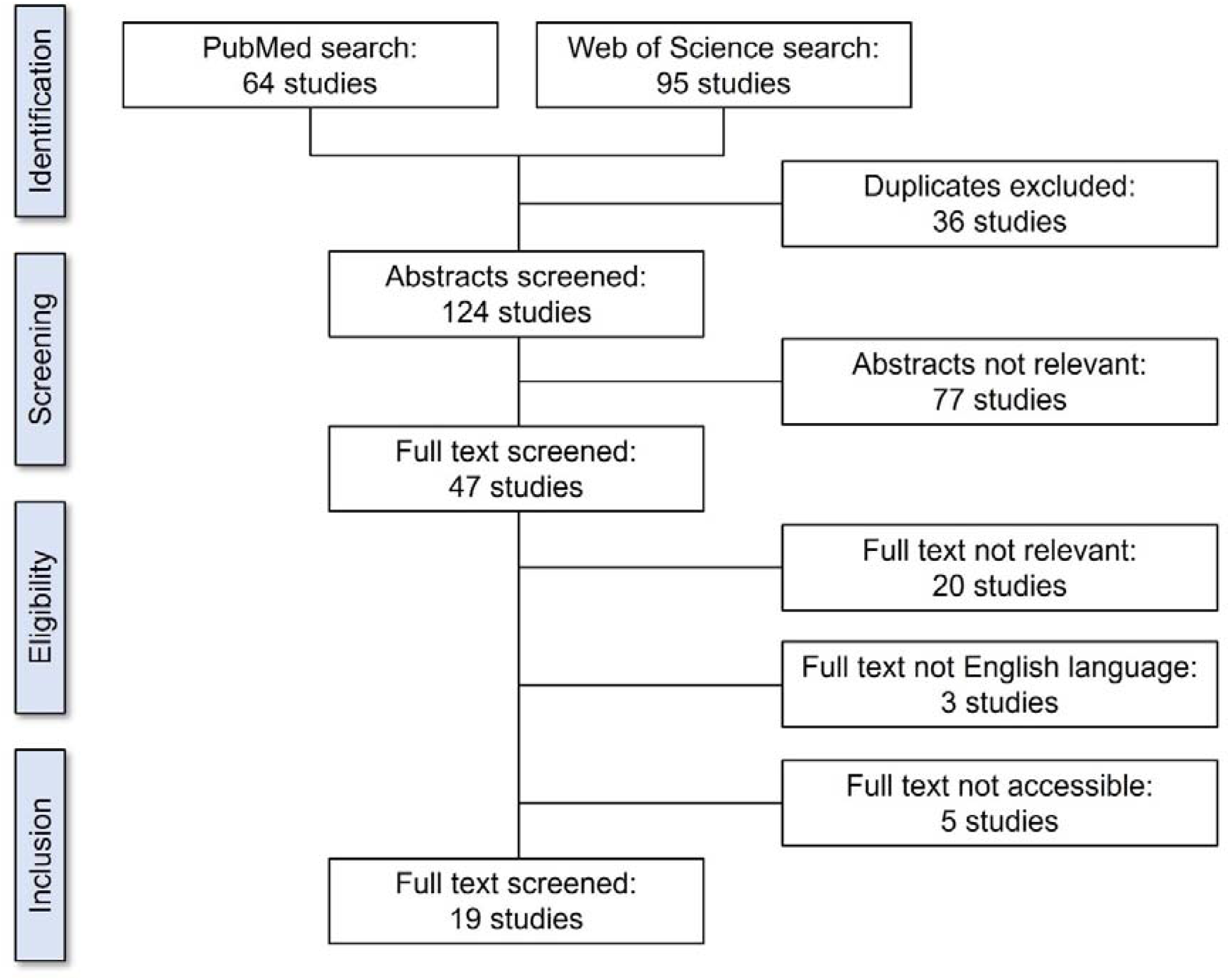
PRISMA 2020 flow diagram for study eligibility.

**Table 1:** Characteristics of eligible studies.

| Ref | Author | Country of study | Funding source | Study design | Study aims | Total participants, N (% female) | Population assessed | Age at baseline | Selection criteria | Outcome(s) | Risk factors / positive associations | Protective factors / negative associations |
| --- | --- | --- | --- | --- | --- | --- | --- | --- | --- | --- | --- | --- |
| [28] | Aarnivala | Finland | Alma and K A Snellman Foundation and The Foundation for Pediatric Research, Finland | Retrospective cohort study | To assess risk factors for the development of DP and torticollis | 155 (51% female) | Healthy newborns | <72 hours from birth | Healthy newborns born between February 2012 and September 2013. Newborns were excluded if having chromosomal anomalies, cleft lip and/or palate, or craniosynostosis | OR/RR for risk factors for DP | - | Risk of developing DP was not associated with torticollis |
| [29] | Aarnivala | Finland | Alma and K A Snellman Foundation | Prospective cohort study | To assess risk factors for cranial deformation by measuring cranial asymmetry from three to 12 months of age using 3D stereophotogrammetry | 99 (47% female) | Healthy newborns | Recruited at birth | Healthy newborns recruited at birth residing within 20 min driving distance from Oulu University Hospital, Finland. Cases of craniosynostosis, cleft lip and/or palate, or syndromic features were excluded | OR/RR for risk factors for DP | DP at six months associated with position preference and imbalance in head rotation at three months. At six months, DP associated with reaching fewer motor milestones by six months. At 12 months, DP associated with position preference at three months and imbalance in head rotation at three months and six months. At 12 months, DP associated with fewer motor milestones and spending more time supine on the floor at six months. Position preference at three months was associated with DP at 12 months. At six months, DP was associated with positional preference at three months and less motor milestones reached by six months. At twelve months, DP was associated with positional preference at three months | Position preference at six months was not associated with DP at 12 months. There no association between DP at 12 months and imbalance in head rotation at three months. No association with DP was also observed for primary sleeping position, time spent in carriers/bouncers/car seats, time spent prone on the floor, pacifier use, illness history (acute otitis media, conditions requiring prolonged hospitalisation), duration of full breastfeeding |
| [30] | Ballardini | Italy | Paediatric Department of the University of Ferrara, Italy | Prospective cohort study | To assess prevalence and risk factors for DP in full term infants | 283 (53% female) | Healthy newborns | Age, weeks (range): All infants, 11.6 (9.4-12.9); infants with DP, 11.7 (10.0-12.8); infants without DP, | Healthy infants born at term presenting at a public immunisation clinic in Ferrara at eight to 12 weeks of age, were included. Infants affected by craniosynostosis, malformations, | OR/RR for risk factors for DP | Maternal/infant risk factors associated with DP included maternal age, supine sleeping position and head position preference | Maternal/infant risk factors not associated with DP included infant sex, maternal origin, maternal education, breast milk feeding, changing crib end, tummy time, instruction about |
|  |  |  |  |  |  |  |  | 11.6 (9.4-12.9) | neurological diseases or admitted to the neonatal intensive care unit (NICU), were excluded |  |  | tummy time |
| [31] | Ifflaender | Germany | The Else Kröner-Fresenius Trust | Cross-sectional study | To assess head shape to determine the prevalence of symmetrical and asymmetrical head deformities and identify possible risk factors | 195 (48% female) | Healthy newborns | Mean postmenstrual age, weeks (standard deviation (SD)): 38.4 (0.9) | Preterm infants discharged from an intermediate care (IMC) unit of a tertiary neonatal clinic in Dresden, Germany from April 2011 to January 2013, were included. All infants were included that were present on the ward at the time of measurement. Those with peripheral cannula at the scalp or requiring supplemental oxygen, were excluded | OR/RR for risk factors for DP | Cranial vault asymmetry index (CVAI ((diagonal A – diagonal B) / diagonal A × 100, where diagonal A > diagonal B)) at term-equivalent age (TEA) was significantly higher (4.1%, interquartile range (IQR) 1.9–6.5%) in very preterm infants compared to late preterm infants and term infants. Moderate or severe DP at TEA was associated with intracranial haemorrhage in preterm infants. Duration of total respiratory support was significantly higher in cases of DP compared to controls. Duration of CPAP therapy was longer in cases of DP compared to controls | No association with DP was observed for sex, bronchopulmonary dysplasia, necrotising enterocolitis, and duration of intermittent mandatory ventilation (IMV) |
| [32] | Kim | South Korea | - | Case-control study | To determine risk factors for DP and its severity, including the association between DP and infant obesity as defined by body mass index (BMI) | 135 infants (40% female) | Infants with cranial deformation on physical exam, with age and sex matched controls randomly selected from the national health screening system for infants and children (confirmed to have no cranial deformation on physical exam) | 2–12 months | Infants two and 12 months old at the time of diagnosis of DP, were included. Infants with neuroimaging results positive for craniosynostosis were excluded. Infants with insufficient clinical data, congenital muscular torticollis, congenital anomalies (e.g. craniofacial/ chromosomal anomalies) or birth injury were also excluded. Age and sex matched controls (confirmed to have no cranial deformation on physical exam) were randomly selected from the national health screening system | OR/RR for risk factors for DP | Factors associated with DP included bottle only feeding, reduced tummy time when awake, delayed motor development, and obesity at diagnosis | Factors not associated with DP included delayed motor development, infant male gender, maternal age, macrocephaly at birth, and macrocephaly, underweight or overweight at the time of diagnosis |
| [33] | Leung | Australia | - | Prospective | To assess association | 94 (59%) | Healthy | Mean age, | Health newborns, born at | Correlation | DP at nine weeks was | No association between |
|  |  |  |  | cohort study | between DP and head orientation or head strength | female) | newborns | days (SD): 21.4 (2.29) | full term (37-42 weeks) between June 2011 and July 2013. Infants were excluded if low birth weight (<2500 g), congenital muscular torticollis, craniosynostosis, neurological insult, or other medical/orthopaedic conditions | between risk factor severity and DP severity | associated with asymmetrical head orientation duration at three weeks and six weeks of age. DP at nine weeks was significantly associated with asymmetrical head orientation strength at three weeks and six weeks of age | DP and latency to turn head |
| [34] | Maniglio, | Italy | - | Prospective cohort study | To assess factors associated with DP to improve screening strategies to identify infants at risk of developing severe deformation | 4377 infants | Infants with DP and controls without DP | 6-12 weeks | Infants born at the San Pietro Fatebenefratelli Hospital in Rome, Italy, during the period from January 2017 to September 2018, were recruited. Infants with congenital deformations, born before 24 weeks gestation, and infants with needed long intensive care treatment, were excluded | OR for risk factors for DP | Maternal age was associated with DP | Male gender was not associated with DP |
| [35] | Mawji | Canada | The Faculty of Graduate Studies, University of Calgary provided \$3000.00 for data collection | Prospective cohort study | To assess potential risk factors for DP in infants seven to 12 weeks of age in Calgary, Alberta | 440 (40.7% female) | Healthy full-term infants | Mean age, weeks: 9 | Healthy full-term infants (born at ≥37 weeks gestation) ranging from seven to 12 weeks of age who presented for immunization at a two-month well-child clinic in Calgary, were included | OR/RR for risk factors for DP | Significant difference in incidence of DP in infants placed supine to sleep compared with sleep in other positions (including prone, side or a combination of supine, prone or side). DP also associated with head positional preference (right and left), maternal education level, and supine sleep position | No association of DP with average length of time in Canada; infant feeding position; and length of tummy time received, male infant sex, and mothers who had a language barrier compared |
| [36] | Nuysink | The Netherlands | - | Prospective cohort study | To assess predictive factors for DP over six months corrected age (CA) in infants | 120 (45.8% female) | Newborns admitted to NICU at gestational age (GA) <30 weeks or birthweight <1000 g | <1 week old | Infants born in, or referred to, a NICU within one week of birth between January 2009 and October 2010. Infants born at GA <30 weeks or birth weight <1000 g who visited the neonatal follow-up clinic, were included. Infants diagnosed with a disease or dysfunction leading to symptomatic asymmetry, such as a central nervous system disorder or congenital malformation, were excluded | OR for risk factors for DP | Association between DP and mechanical ventilation and chronic lung disease grade II in the neonatal period | - |
| [37] | Nuysink | Finland | The University of Oulu Scholarship Foundation, the Orthodontic Section of the Finnish Dental Association Apollonia, the Emil Aaltonen Foundation, the Alma, and KA Snellman Foundation, the Finnish Medical Foundation, and the Foundation for Pediatric Research in Finland | Case-control study | To utilise 3D stereophotogrammetry to assess cranial growth, moulding, and incidence of DP in preterm children compared to term born children | 68 (32% female) | Healthy newborns | Gestational age, weeks: Preterm, 32.7; term, 40.0 | Infants were considered eligible if they had no cheilopalatoschisis, craniosynostosis, or dysmorphic features and if they resided within Oulu region, Finland. All participants were born between 2012 and 2015 in Oulu University Hospital. The control group was randomly selected from a previously collected non-intervention cohort by computer-based random selection and matched to gender | Oblique cranial length ratio (OCLR) mean difference | - | No difference in head shape (OCLR) between preterm and full-term children or between genders |
| [38] | Pogliani | Italy | - | Retrospective cohort study | To assess risk factors for DP at birth | 413 (50% female) | Healthy newborns | <72 hours from birth | Neonates >33 weeks GA born between May 2011 and January 2012. Newborns with extreme low birth weight or presenting with major congenital malformations needing NICU transfer were excluded. Children of mothers with documented TORCH infection were not recruited | OR/RR for risk factors for DP | Association between DP and male gender | - |
| [39] | Roberts | UK | - | Retrospective cohort study | To test the hypothesis that ventriculoperitoneal (VP) shunt insertion significantly increases contralateral DP | 339 (42% female) | Children aged 0–16 years with VP shunts | - | Children aged 0–16 years with at least one follow-up scan from surgical database at Paediatric Neurosurgery, Birmingham Children's Hospital between 2006 and 2013. Children without imaging were excluded | OR/RR for risk factors for DP | DP is associated with occipital shunt placement. A statistically significant difference in probability of becoming plagiocephalic between neonates and 12–16 year olds was observed. Boys were more likely to develop shunt-associated plagiocephaly than girls | No difference in DP between neonates and infants, neonates and those aged 3–5 years, neonates and 1–3 year olds and between neonates and 5–12 year olds |
| [40] | Sheu | US | Cooperative agreement U01DD000494 from the Centers for Disease Control and Prevention and | Retrospective cohort study | To assess factors that may explain a 9-fold increase in plagiocephaly in Texas from 1999 to 2007 | 6295 infants (38% female) | Infants with DP | - | Cases identified using the Texas Birth Defects Registry (TBDR) with a definite diagnosis of DP (BPA code 754.050), born from January 1, 1999, through December | Mean difference/percentage difference for risk factors for DP | Lower maternal education was associated with DP | No association of DP with maternal age or race/ethnicity, infant sex. Mean age at DP diagnosis did not significantly change over time |
|  |  |  | Title V Maternal and Child Health Block Grant funds from the Health Resources and Services Administration |  |  |  |  |  | 31, 2007. Cases of plagiocephaly with craniosynostosis were excluded |  |  |  |
| [41] | Solani | Iran | Grant funding from the Vice-chancellor for Research, KAUMS, Kashan, Iran | Case-control study | To determine the risk factors of positional plagiocephaly in healthy infants | 300 infants | Healthy Iranian infants | 8–12 weeks | Healthy full-term infants (gestational age >37 weeks) aged 8–12 weeks who were referred to the paediatric neurology clinic at Shahid Beheshti Hospital (Kashan, Iran) affiliated with Kashan University of Medical Sciences (KAUMS), were included | OR for risk factors for DP | Factors associated with DP included male gender, head circumference and supine sleeping position | Firmness of headrest was not associated with DP |
| [42] | Tang | USA | - | Prospective cohort study | To assess prevalence of DP in infants with neonatal brachial plexus palsy (NBPP) and spontaneous recovery from DP | 28 (50% female) | Full-term infants >3 months and <1 year of age with NBPP | Mean age, months (SD): 3 (3) | Full-term infants >3 months and <1 year of age with NBPP. Patients with neurological or congenital comorbidities in addition to NBPP, helmet therapy for plagiocephaly, and/or surgical procedures related to NBPP were excluded. Children with craniosynostosis were excluded | Mean difference/percentage difference for risk factors for DP | - | Maternal age and race (white/black) were not associated with DP |
| [43] | Valkama | Finland | - | Case-control study | To assess the prevalence of DP in children with developmental dysplasia of the hip (DDH) | 120 (56% female) | Children with DDH with or without deformational plagiocephaly | Age, years (SD): DDH children, 8.0 (1.4); matched controls, 7.9 (1.3) | DDH cases from newborn infants born at the University Hospital, University of Oulu, Finland, were included. Preterm and disabled children were excluded | OR/RR for risk factors for DP | 10.3% of DDH children and only 1.5% of control children had DP | OCLR was equal between DDH children and controls. No association between side of DDH and DP |
| [44] | van Vlimmeren | The Netherlands | Grant BU002/10 from the Scientific Committee of The Royal Dutch Association for Physiotherapy, Amersfoort, The Netherlands | Prospective cohort study | To assess skull shape in healthy newborns until the age of 5.5 years, in children with and without positional preference at 7 weeks and in the children with positional preference who received paediatric physical therapy intervention (PPT) or no PPT | 380 (53% female) | Healthy newborns | <48 hours from birth | Healthy newborns (≥36-weeks gestation), born between December 2004 and September 2005 at the general district hospital Bernhoven in Veghel, The Netherlands. Children with congenital muscular torticollis (Kaplan type 2 and 3), dysmorphism or syndromes, were excluded | Mean difference/percentage difference for risk factors for DP | Association between DP and position preference | No association between potential risk factors (nursing, feeding, sleeping, and playing positioning habits) at seven weeks of age and skull deformity at 24 months and 5.5 years of age. A trend towards significance between time spent playing prone ('tummy time') at seven weeks of age and the ODDI percentage at 24 months of age |
| [45] | Weernink | The | ZonMw, the | Case-control | To assess the influence | 823 infants | Infants with and | 2–4 months | Children born between | OR/RR for | Insufficient vitamin D | Maternal |
|  |  | Netherlands | Netherlands Organization for Health Research and Development (grant number 170.992.501) | study | of adherence to recommendations for vitamin D supplement intake of 10 µg per day (400 IU) in the first months of life (child) on the occurrence of DP of the child at the age of 2 to 4 months | (46% female) | without DP | old | November 22, 2009 and June 9, 2010, with mild to severe DP from Helmet Therapy Assessment in Deformed Skulls (HEADS) study. Controls were included from a 2010 survey on infant milk feeding | risk factors for DP | supplement intake during early infancy was associated with DP. Maternal sociodemographic factors associated with DP included mother's age and mother's education level. Infant factors associated with DP included male gender, formula feeding, and milk formula consumption after birth | sociodemographic factors not significantly associated with DP included mother's country of birth (other than Netherlands). Infant factors not associated with DP included time child spent outdoors |
| [9] | van Cruchten | The Netherlands | - | Cohort study | To assess the impact of risk factors on the type and severity of DP | 184 infants (30.4% female) | Infants seen in outpatient clinic with a parental concern for DP | 3–14 months old | Exclusion criteria consisted of children above the age of 14 months or below 3 months of age and other forms of cranial deformations, such as cranial synostosis | Differences in ODDI for risk factors for DP | Negative correlation between age and ODDI. Positive correlation between ODDI and position preference right and position preference left | No association between ODDI and developmental delay, family history of DP, gender, and torticollis |

### Demographic factors

*Age:* One study reported a significant negative correlation between age and oblique diameter difference index (a biomarker for plagiocephaly) [9]. However, a second study (which assessed factors associated with a nine-fold increase in plagiocephaly between 1999 to 2007 in Texas) reported no association between age and DP [40]. Finally, a third study (which assessed DP in children with VP shunts) reported that infants were more likely to develop shunt-associated DP compared to older children, although no statistical test of significance for this association was reported [39].

*Gender:* Four studies demonstrated a significant association between male gender and DP [35, 38, 41, 45], one reported borderline significance [39], and seven reported no association between male gender and DP [9, 30–32, 34, 37, 40]. Of these 12 studies, six studies [31, 32, 35, 38, 41, 45] reported odds ratios which were analysed by meta-analysis, which revealed significant inter-study heterogeneity (I^2^, 68.5%; Q, 15.89; df, 6). Pooled effects revealed fixed and random effects for OR for DP for male gender of 1.71 (95% CI 1.43-2.04; P<0.001) and 1.51 (95% CI 1.07-2.12; P=0.017), respectively (Figure 2A). A funnel plot with asymmetry analysis excluded publication bias (P=0.1201) (Figure 2B).

**Figure 2:**
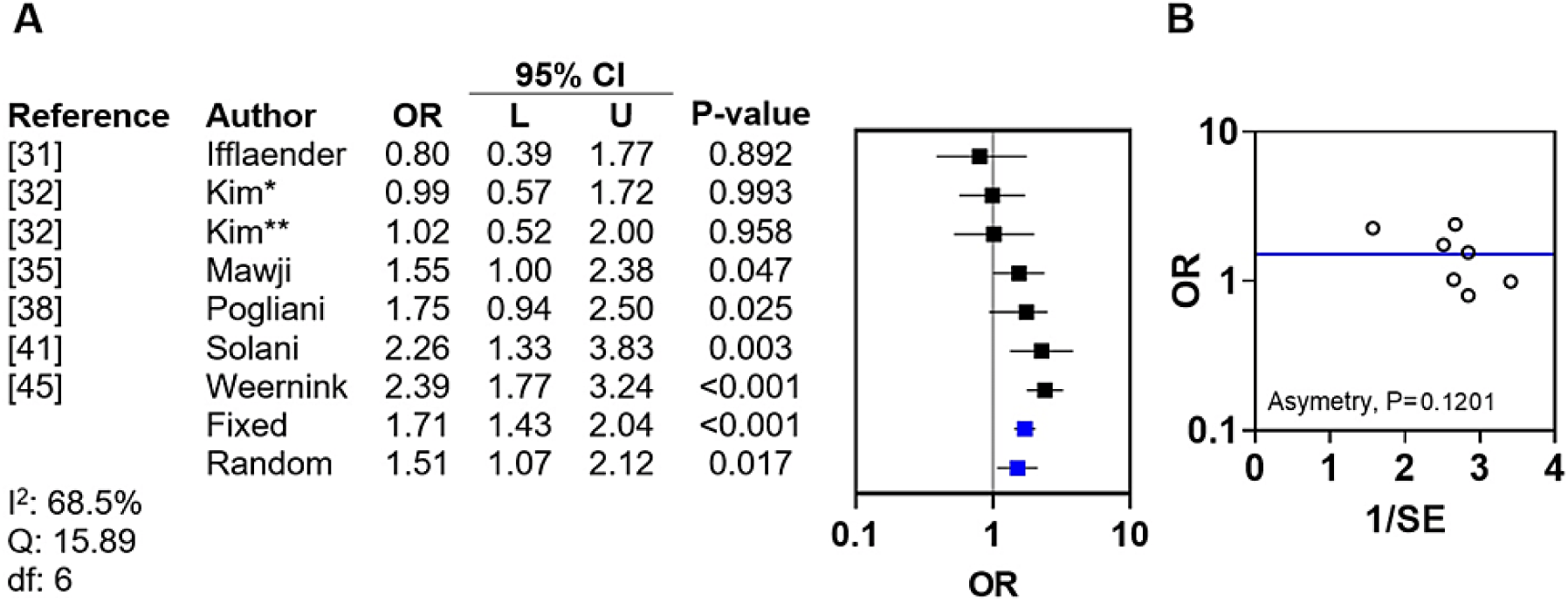
Meta-analysis and funnel plot for male gender. A) Forrest plot of OR for DP for male gender with fixed and random effects; B) funnel plot with linear regression test of asymmetry; I^2^, inconsistency (heterogeneity) index; Q, chi-squared significance; df, degrees of freedom; 95% CI, 95% confidence interval; L, lower limit of the 95% CI; U, upper limit of the 95% CI; *, mild to moderate DP (univariate analysis); **, severe DP (univariate analysis); blue line, random effects.

*Race:* Race was investigated by one study, which included a cohort of children with brachial plexus palsy [42]. In this group, race was not significantly associated with DP that developed following brachial plexus injury.

### Developmental factors

*Developmental delay:* Developmental delay was investigated as a risk factor for DP by one study, which concluded developmental delay was not associated with DP [9].

*DDH:* DDH was investigated as a risk factor for DP by one study, which assessed the prevalence of DP in children with DDH [43]. DDH was significantly associated with DP compared to controls without DDH.

*Head circumference:* Head circumference was investigated by one study, which concluded head circumference was associated with an OR for DP of 1.22 (95% CI 1.06-1.40) [41].

*Motor milestones:* Reaching fewer motor milestones by six months of age was investigated as a risk factor by two studies [29, 32]. The first study reported that reaching fewer motor milestones by six months was associated with an adjusted OR for DP of 2.35 (95 % CI 1.25-4.42) [29]. The second study found an association between delay in motor development and DP [32]. Odds ratios from these studies were analysed by meta-analysis, which identified no inter-study heterogeneity (I^2^, 0.0%; Q, 1.67; df, 3) (Figure 3A). Pooled fixed and random effects for OR for DP for delayed motor milestones were 2.56 (95% CI 1.66-3.96, P<0.001) and 2.56 (95% CI 1.66-3.96, P<0.001), respectively. A funnel plot with asymmetry analysis excluded publication bias (P=0.1829) (Figure 3B).

**Figure 3:**
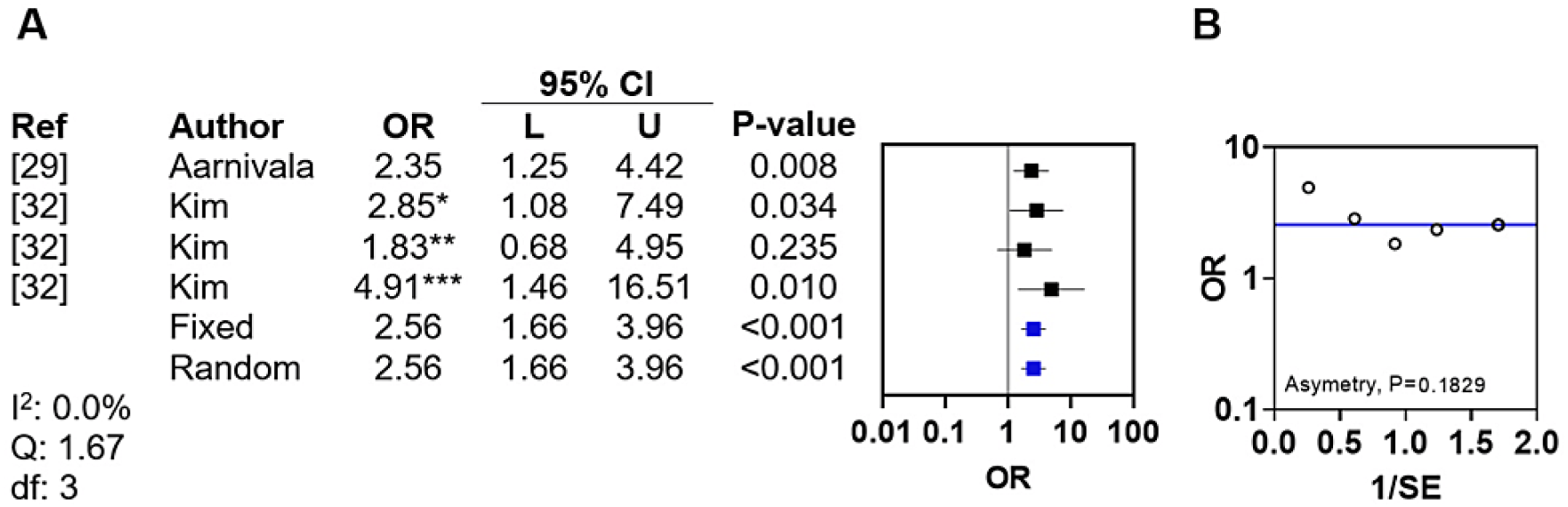
Meta-analysis and funnel plot for reaching fewer motor milestones by six months of age. A) Forrest plot of OR for DP for reaching fewer motor milestones by six months of age with fixed and random effects; B) funnel plot with linear regression test of asymmetry; I^2^, inconsistency (heterogeneity) index; Q, chi squared significance, df, degrees of freedom, 95% CI, 95% confidence interval, L, lower limit of the 95% CI; U, upper limit of the 95% CI;*aOR for DP (multivariate analysis),**OR mild to moderate DP (only univariate analysis available),***aOR for Severe DP (multivariate analysis); blue line, fixed effects

*Overweight/underweight:* Being overweight at diagnosis of DP was investigated as a risk factor for DP by one study, which reported being overweight at diagnosis was not significantly associated with DP [32]. The same study reported that being underweight at diagnosis was also not associated with DP.

### Dietary factors

*Bottle only feeding:* Bottle only feeding was investigated by one study, which demonstrated bottle only feeding was associated with an adjusted OR for DP of 4.65 (95% CI 2.70-8.00) [32].

*Breast feeding:* Duration of exclusive breast feeding was investigated by one study, which demonstrated that duration of exclusive breast feeding was not significantly associated with DP [29].

*Formula feeding:* Formula feeding was investigated by one study [45]. This study reported that children that developed DP by two to four months of age were significantly more likely to be formula fed (adjusted OR, 1.51; 95% CI 1.00-2.27) [45].

*Vitamin D intake:* Vitamin D intake in the infant was investigated by one study [45]. This study reported that children that developed DP by two to four months of age were significantly more likely to have insufficient vitamin D intake (adjusted OR, 7.15; 95% CI 3.77-13.54) [45].

### Maternal factors

*Maternal age:* Three studies demonstrated an association between maternal age and DP [30, 34, 45]. Of these three, one study reported that mothers of infants with a diagnosis of DP were older than mothers of infants without DP (36.2; 95% CI 27.04-45.37 vs 34.0; 95% CI 22.89-45.12, respectively; P<0.01) [34]. Three further studies reported no significant association [32, 40, 42]. Only one of the six studies reported an OR, which suggested increased maternal age was a protective factor for the development of DP (OR, 0.94; 95% CI 0.91-0.97) [45].

*Maternal education level:* Three studies demonstrated a significant association between maternal education level and DP [35, 40, 45] and one reported no significant association [30]. Only two studies reported odds ratios [35, 45]. Of these two, one study indicated that low education level was an adverse factor in the development of DP (adjusted OR, 1.97; 95% CI 1.19-3.26) [45], while the other study suggested postsecondary education was not a significant protective factor (OR, 0.71; 95% CI 0.43-1.16) [35]. The OR from the former was utilised in a meta-analysis with the inverse OR from the latter study, which demonstrated no inter-study heterogeneity (I^2^, 0.0%; Q, 0.86; df, 1) (Figure 4). The pooled effects for OR for DP for low education level revealed a fixed effects of 1.66 (95% CI 1.17-2.37, P<0.005).

**Figure 4:**
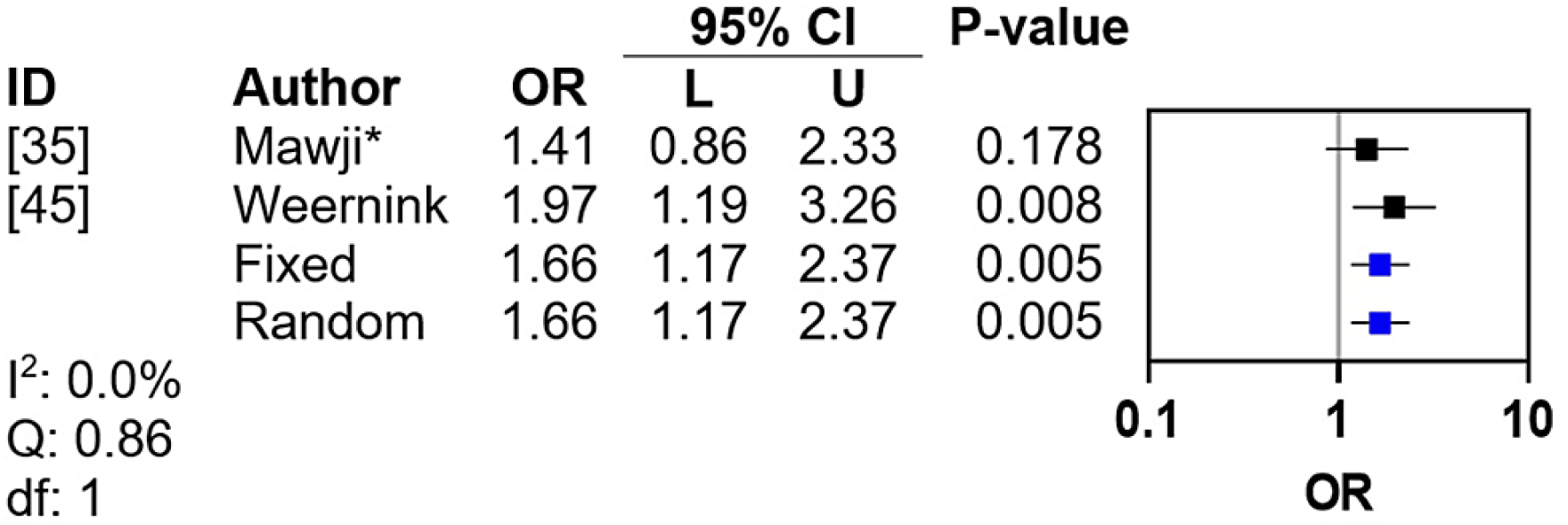
Meta-analysis for maternal education level. I^2^, inconsistency (heterogeneity) index; Q, chi squared significance, df, degrees of freedom, 95% CI, 95% confidence interval, L, lower limit of the 95% CI; U, upper limit of the 95% CI; *, reciprocal OR for postsecondary education

*Maternal language barriers:* One study investigated whether infants with mothers who experience a language barrier when receiving medical advice had a higher rate of DP [30]. This study suggested maternal language barrier was not associated with DP in the infant.

*Maternal race/country of origin:* Three studies investigated whether maternal race/country of origin [30, 40, 45]. All three studies suggested maternal race/country of origin was not associated with DP in the infant.

*Length of time in country of study:* One study investigated whether length of time spent by mothers in the country in which the study was conducted was associated with DP [35]. This study reported length of time in country of study was not associated with DP.

*Pacifier use:* Pacifier use by mothers in healthy infants was investigated as a risk factor for DP by one study, reported pacifier use was not associated with DP [29].

*Tummy time instructions:* One study investigated whether DP was associated with mothers receiving instructions about tummy time [30]. This study reported that receiving instructions about tummy time was not associated with DP.

### Medical and surgical factors

*Bronchopulmonary dysplasia:* Bronchopulmonary dysplasia was investigated as a risk factor for DP by one study, which included a cohort of infants born prematurely [31]. Bronchopulmonary dysplasia was not associated with DP [31].

Chronic lung disease: Chronic lung disease grade II was investigated by one study, which also assessed DP risk factors in infants born prematurely [36]. Chronic lung disease grade II was reported to be significantly associated with DP [36].

*Family history of DP:* Family history of DP was investigated by one study that suggested family history of DP was not associated with DP [9].

*History of illness:* History of illness was investigated by one study which suggested history of illness was not associated with DP [29].

*Intracranial haemorrhage:* Intracranial haemorrhage was investigated by one study, which assessed DP risk factors in infants born prematurely [31]. Intracranial haemorrhage was not associated with DP [31].

*Macrocephaly:* Macrocephaly at birth was investigated by one study [32]. This study suggested macrocephaly at birth was not associated with DP. Macrocephaly at diagnosis of DP was also investigated, but also was not associated with DP (OR 1.38; 95% CI 0.63-3.04) [32]. Lack of association was reported for subgroups with mild to moderate DP (OR 1.48; 95% CI 0.62-3.53) and severe DP (OR 1.19; 95% CI 0.04-3.58) [32].

*Mechanical ventilation:* Two studies demonstrated an association between mechanical ventilation and the development of DP in preterm infants [31, 36]. Of these, one study reported an OR for mechanical ventilation (OR 1.10; 95% CI 1.00-1.14) [36]. The other study also suggested duration of total respiratory support (continuous positive airway pressure (CPAP) and intermittent mandatory ventilation (IMV)), and duration of CPAP alone, were associated with DP, while IMV alone was reported not associated with DP [31].

*Necrotising enterocolitis:* Necrotising enterocolitis was investigated by one study, which included a cohort of infants born prematurely [31]. Necrotising enterocolitis was not associated with DP [31].

*Obesity:* Obesity at diagnosis of DP was investigated by one study [32]. Obesity at diagnosis of DP was defined as BMI >97th percentile. This study concluded that obesity at diagnosis of DP was associated with DP (adjusted OR, 2.45 95% CI 1.02-5.90) [32]. This study also suggested obesity at diagnosis of DP was more significant in severe DP (adjusted OR 4.10, 95% CI 1.42-11.90), but not associated with mild to moderate DP (adjusted OR 2.29; 95% CI 0.86-6.05).

*Occipital shunt placement:* Occipital shunt placement was investigated by one study, which concluded occipital shunt placement was not associated with DP [31].

*Torticollis:* Torticollis was investigated by three studies, all of which reported no association between torticollis and DP [9, 28, 40].

### Positional and environmental factors

*Carriers/bouncers/car seats/headrests:* Time spent in carriers/bouncers/car seats was investigated by one study [29]. This study reported time spent in carriers/bouncers/car seats was not associated with DP. Firmness of headrest was investigated by another study [41]. This study reported firmness of headrest was not associated with DP (OR, 1.31; 95% CI 0.72-2.37) [41].

*Change of crib end:* Change of crib end was investigated as a risk factor for DP by two studies that suggested change of crib was not associated with DP [35, 44].

*Feeding position:* Feeding position was investigated as a risk factor for DP by two studies that demonstrated feeding position was not associated with DP [35, 44].

*Latency in head turning:* Latency in head turning was investigated by one study, which concluded latency in head turning was not associated with DP [33].

*Playing position:* Playing position was investigated by one study, which demonstrated playing position was not associated with DP [44].

*Position preference:* Position preference was investigated by six studies, all of which demonstrated an association between head position preference and DP [9, 29, 30, 33, 35, 44]. Of these, two studies [35, 36] reported ORs which were utilised in meta-analysis that revealed no significant inter-study heterogeneity (I^2^, 27.6%; Q, 2.76; df, 3) (Figure 5A). Pooled effects for OR for DP for head position preference for fixed-and random-effects were 4.75 (95% CI 3.36-6.73, P<0.001) and 4.96 (95% CI 3.10-7.93, P<0.001), respectively. A funnel plot with asymmetry analysis excluded publication bias (P=0.2849) (Figure 5B).

**Figure 5:**
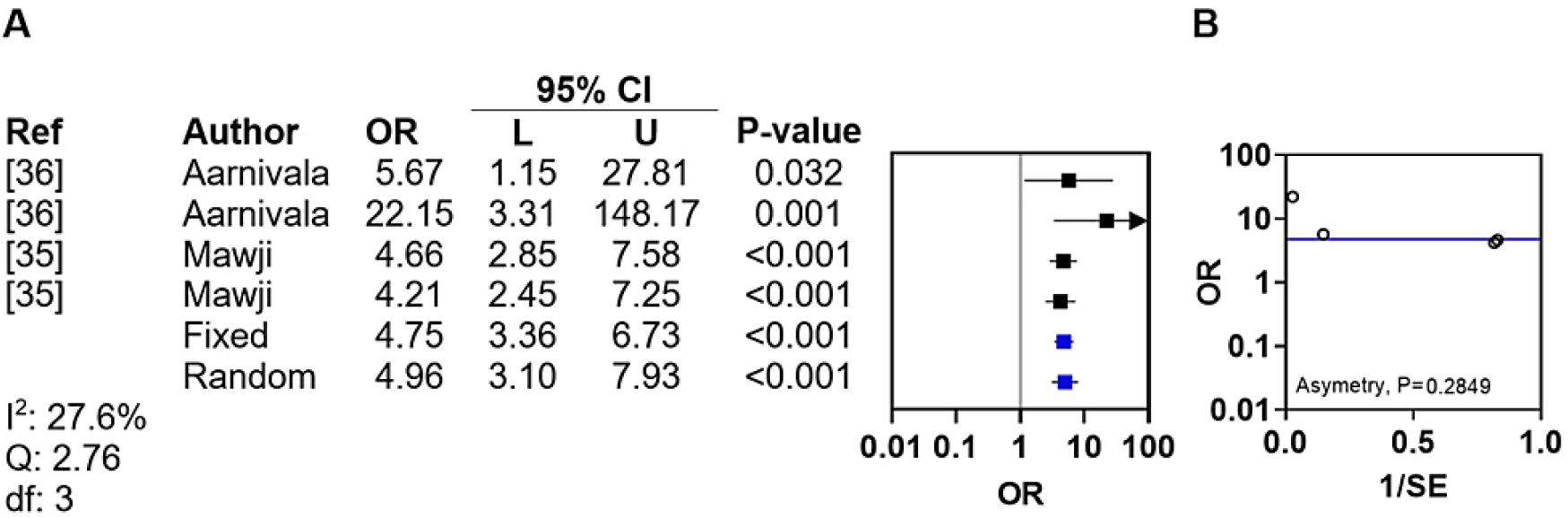
Meta-analysis and funnel plot for head position preference. A) Forrest plot of OR for DP for head position preference with fixed and random effects; B) funnel plot with linear regression test of asymmetry; I^2^, inconsistency (heterogeneity) index; Q, chi squared significance; df, degrees of freedom; 95% CI, 95% confidence interval; L, lower limit of the 95% CI; U, upper limit of the 95% CI.

*Sleeping position:* Sleeping position (supine vs prone) was investigated by three studies [30, 35, 41]. The first two studies demonstrated an association between sleeping supine and DP [35, 41]. The third study reported a protective effect from sleeping prone (OR, 0.13; 95% CI 0.03-0.40) or side sleeping (OR, 0.22; 95% CI 0.05-0.71) [30]. The ORs from the first two studies were utilised with the inverse OR from the third study in a meta-analysis, which demonstrated no significant inter-study heterogeneity (I^2^, 19.1%; Q, 2.47; df, 3) (Figure 6A). The pooled effects for OR for DP for sleeping position revealed fixed-and random-effects of 3.12 (95% CI 2.21-4.39, P<0.001) and 3.22 (95% CI 2.14-4.84, P<0.001), respectively. A funnel plot with asymmetry analysis excluded publication bias (P=0.2021) (Figure 6B).

**Figure 6:**
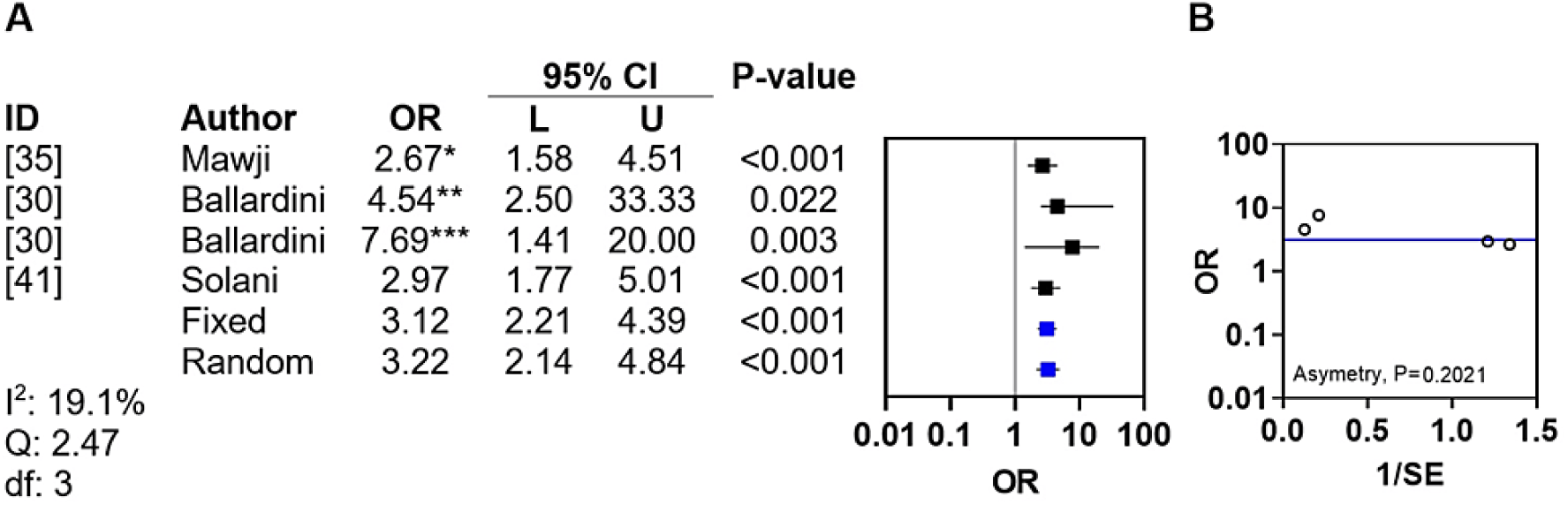
Meta-analysis and funnel plot for supine sleeping position. A) Forrest plot of OR for DP for supine sleeping position with fixed and random effects; B) funnel plot with linear regression test of asymmetry; I^2^, inconsistency (heterogeneity) index; Q, chi squared significance, df, degrees of freedom, 95% CI, 95% confidence interval, L, lower limit of the 95% CI; U, upper limit of the 95% CI;*Supine vs Prone, **Reciprocal prone vs supine, ***Reciprocal side vs supine

*Time spent prone on the floor:* One study investigated time spent prone on the floor and reported no associated with DP [29].

*Tummy time:* Tummy time was investigated by four studies [30, 32, 35, 44]. One study reported an association between reduced tummy time when awake and DP (aOR, 3.51; 95% CI 1.71-7.21) [32]. The remaining studies reported that tummy time was not associated with DP, but provided no ORs [30, 35, 44].

*Time spent outdoors:* Time spent outdoors was investigated by one study, which reported no association with DP [45].

*Time spent supine on the floor:* Time spent supine on the floor was investigated as a risk factor for DP by one study [29]. The study reported that children with DP at 12 months spent more time supine on the floor at six months [29].

### Summary of non-obstetric factors associated with DP

A summary of ORs (where available) for non-obstetric factors significantly associated with DP, is illustrated in Table *2*.

**Table 2:** Odds ratios for factors associated with DP.

| Factor | OR | 95% CI | Ref |
| --- | --- | --- | --- |
| Insufficient vitamin D intake | 7.15 | 3.77–13.54 | [45] |
| Head position preference | 4.75 | 3.36–6.73 | [35, 36] |
| Bottle only feeding | 4.65 | 2.70–8.00 | [32] |
| Reduced tummy time | 3.51 | 1.71–7.21 | [32] |
| Sleeping position | 3.12 | 2.21–4.39 | [30, 35, 41] |
| Fewer motor milestones by six months of age | 2.56 | 1.66–3.96 | [29, 32] |
| Obesity (BMI >97<sup>th</sup> percentile) | 2.45 | 1.02–5.90 | [32] |
| Maternal education level | 1.66 | 1.17–2.37 | [35, 45] |
| Male gender | 1.51 | 1.07–2.12 | [31, 32, 35, 38, 45] |
| Formula feeding | 1.51 | 1.00–2.27 | [45] |
| Macrocephaly at diagnosis | 1.38 | 0.63–3.04 | [32] |
| Head circumference | 1.22 | 1.06–1.40 | [41] |
| Mechanical ventilation | 1.10 | 1.00–1.14 | [36] |
| Chronic lung disease grade II | - | - | [36] |
| DDH | - | - | [43] |
| Maternal age | - | - | [30, 34, 45] |
| Time spent supine | - | - | [29] |

## Discussion

This study assessed evidence of association with DP for 43 non-obstetric factors. This included three demographic factors, six developmental factors, four dietary factors, eight maternal factors, 11 medical and surgical factors, and 11 positional and environmental factors. Of these, a total of 17 factors were associated with DP. This included one demographic factor (male gender), three developmental factors (DDH, head circumference, and delay in motor milestones), three dietary factors (bottle only feeding, formula feeding and vitamin D intake), two maternal factors (maternal age and maternal education level), three medical and surgical factors (chronic lung disease grade II, macrocephaly, mechanical ventilation and obesity at diagnosis), and four positional and environmental factors (head position preference, sleeping position, reduced tummy time, and time spent supine on the floor). With the notable exceptions of maternal age, mechanical ventilation and tummy time, these associations were either supported by non-conflicting evidence or a meta-analysis that resolved conflicting evidence into a significant association. Thirteen factors had significant ORs that ranged from 1.10 (mechanical ventilation) to 7.15 (insufficient vitamin D intake). Of the five factors assessed by meta-analysis (male gender, reaching fewer motor milestones by six months of age, maternal education level, head position preference, sleeping position), only one (male gender) was associated with significant inter-study heterogeneity. No evidence of publication bias was detected.

### Evidence-based recommendations

Strategies to reduce the prevalence of DP have included guidance about the infant’s environment, positioning, and handling, with the goal of creating a non-restrictive environment that promotes spontaneous and unhindered physical movement and symmetrical motor development [29, 46].

Based on the evidence presented in this study, the following recommendations can be made with the aim of reducing the prevalence of DP. In order of importance, these recommendations include:

To ensure bone health in infants, it is critical that vitamin D intake is adequate [47]. Vitamin D level should be assessed regularly during development and dietary supplementation should be considered if vitamin D level is low.

During the first months of life, babies develop a head position preference [48] and this preference is more often to the right [49, 50]. The increased compressive forces on one side of the head for prolonged periods causes flattening on the side being compressed. It has been proposed that the position of the foetus in the latter stages of pregnancy may, in part, be responsible for positional preference [30, 51].

However, there is also evidence that position preference can be modified by varying head position during sleep, to encourage equal distribution of pressure [52]. Other strategies to mitigate head position preference should be utilised, such as gently moving the infants head to the unfavoured side when asleep and/or physical therapy with or without kinesiological tape to reduce tightness in the neck muscles to facilitate easier neck turning [53, 54]. Sufficient tummy time should be provided to strengthen the infant’s head, neck and arms and reduce time spent supine when awake. Since infants will turn away from windows and towards the centre of a room, the sleeping position should be alternated by placing the infants head at alternate ends of the crib. This concept is supported by two studies suggesting no association between crib end change and DP [35, 44]. It is important however to state that the lack of association between DP and change of crib end does not necessarily mean that changing crib end will provide an effective treatment of established DP, it may merely help to reduce the risk of DP developing.

Although bottle only feeding should not be discouraged, it is recommended this task is performed while alternating feeding positions using both the dominant and non-dominant sides when holding the infant. Bottle feeding, as opposed to exclusively breast feeding, may be positively associated with obesity (which has also been identified as a risk factor for DP) [55]. This co-association may, in part, explain the association of bottle only feeding with DP. Although formula feeding should also not be discouraged, it is recommended this task is performed by alternating feeding positions. This particular risk factor has been suggested by Weernink et al. to be associated with lower maternal education level [45]. Similarly to bottle feeding, formula feeding, as opposed to exclusively breast feeding, may be positively associated with obesity (itself a risk factor for DP) [55].

If motor milestones are delayed, infants should be referred for specialist assessment by a paediatrician. Similarly, obesity (BMI >97th percentile) should be identified and managed by a specialist paediatrician.

Lower maternal education level results in worse health outcomes in infancy and later life [56]. Although improving the education level of future mothers should be a national priority, there are other factors associated with low education level which may be immediately modifiable. These include access to information and access to resources [57]. To mitigate the impact of low maternal education level, educational resources and face-to-face education outlining factors associated with DP, and recommendations to reduce their impact, should be provided to all families at the six-week post-natal infant check-up, but particularly those with low educational levels.

Losee et al. suggested male infants tend to have larger, less flexible heads at birth, that are more likely to become deformed by compressive forces in utero and in the birth canal [51]. Information should be provided to families that male infants are at higher risk of DP. Families should be particularly encouraged to engage with strategies to mitigate DP in male infants. Information should be provided to families that infants with macrocephaly / greater head circumference are at higher risk of DP. Families should be particularly encouraged to engage with strategies to mitigate DP in these infants. Information should also be provided to families that infants who have had mechanical ventilation are at higher risk of DP. Families and care teams for patients requiring mechanical ventilation should be encouraged to engage with strategies to mitigate DP in these infants.

Adopting these recommendations may lead to a reduction in the prevalence of DP. It has been demonstrated that education provided by healthcare professionals, such as health visitors, midwives and nurses, can successfully reduce the prevalence of DP [58]. Alternative strategies for mitigating the impact of DP, such as helmeting therapy, are costly to individuals and healthcare systems and may be associated with morbidity [59].

### Strengths of this study

The strengths of this study include robust methodology. The protocol was registered with PROSPERO and reporting was in-line with PRISMA 2020 guidelines [24]. The databases searched provided greater than 97.5% coverage [25]. Screening required independent reviewers. Standardised data extraction was performed to minimise errors. Meta-analysis was performed where data allowed. Funnel plots suggested publication bias did not impact on the results, thus trim and fill analysis was not necessary to correct for asymmetry [60].

This study represents the most comprehensive analysis of non-obstetric factors associated with DP published to date. A systematic review by Bialocerkowski et al. only identified five factors associated with DP, of which four were non-obstetric factors (male gender, supine position, and neck problems) [61]. Another systematic review by De Bock et al. identified male gender, supine sleep position, limited neck rotation, head position preference, lower level of activity and reduced tummy time, as the most important risk factors [62]. In contrast, our study provides evidence of association for 17 factors, and confirms association of male gender, supine sleep position, head position preference and reduced tummy time.

A more recent systematic review by Inchingolo et al. only provided the following two recommendations to mitigate the impact of non-obstetric risk factors for DP: at least 30 min in tummy time and use of a passive sleep curve mattress to improve harmonious skull growth [63]. It also did not include meta-analyses upon which to base recommendations. In contrast, our study makes 13 recommendations for non-obstetric factors to reduce the prevalence of DP, of which five are based on meta-analytic methods. The systematic review by Inchingolo et al. also did not assess publication bias. Our study suggested that none of the recommendations were influenced by publication bias.

### Limitations of this study

To ensure completion of the study, additional databases, hand searching and grey literature searching was not performed. This inevitably limited the comprehensiveness of this study, although no systematic review can claim to truly be comprehensive as this would necessitate continuous inclusion of newly published studies. Search terms were designed to be specific yet pragmatic; thus some studies may have escaped the searches. Due to resource limitations, it was not possible to record more detailed reasons for exclusion of screened articles other than not reporting risk factors for DP.

These factors may, in part, account for apparent disparities reported in our results compared to other literature. These disparities include the apparent lack of association between developmental delay and DP [9], while demonstrating a significant association between reaching fewer motor milestones and DP [29, 32]. A recent systematic review by Martinuik et al. included 19 studies that assessed an association between developmental delay and DP. Notwithstanding the fact that they included multiple studies that utilised the same study population more than once, a positive association between DP and developmental delay was reported in a majority of studies [64]. The fact that our study did not demonstrate a similar association, may in part be due to inclusion of a study that assessed a different study population, but may also be due to methodological limitations of our search strategy that limited the identification of studies that may have met our study eligibility criteria. Another study of a large primary care cohort of 77,108 patients has provided further evidence in support of an association between DP and developmental delay [65]. That said, the literature remains conflicting, with other studies unable to demonstrate an association between DP and presence or degree of developmental delay [66]. Commentators have highlighted the fact that most studies are retrospective and/or observational by design and this limits conclusions about the correlative versus causative relationship between DP and developmental delay [67].

Another disparity reported in our study is that torticollis was not associated with DP [9, 28, 40]. A number of literature reviews have previously reported an association between congenital muscular torticollis and DP [8, 61]. Although these studies were published a decade or so ago, it logically follows that if other positional factors, such as head position preference and sleeping position, have been found in more contemporaneous studies to be significantly associated with DP [30, 35, 36, 41], then torticollis would also be expected to be found to be associated with DP. These conflicts reported in the literature may be due to limitations of individual study design or differences between different study populations. We therefore recommend that the lack of association between torticollis and DP that is reported in our study should be interpreted with caution, given the lack of a mechanistic explanation that reconciles this result with other positional factors that were found to be associated with DP.

## Conclusions

In summary, this study provides the most comprehensive meta-analytic assessment of non-obstetric factors associated with DP published to date. It provides 13 evidence-based recommendations which can be adopted by healthcare systems globally, to reduce the prevalence of DP and its impact on child development. Future research should focus on investigating factors for which literature is conflicting, but quantitative data is lacking, to enable meta-analyses to be performed. Maternal age was the only factor reported to be protective against DP, although conflicting studies reported no association without reporting quantitative data. Thus, maternal age as a protective factor for DP should be investigated further to provide quantitative data for meta-analytic approaches to determine its protective effect. Finally, randomised controlled trials (RCTs) should be conducted focusing on the effect of the recommendations set out in this study at reducing the prevalence of DP.

## Data Availability

All data produced in the present work are contained in the manuscript

## References

1. Ehret FW, Whelan MF, Ellenbogen RG, Cunningham ML, Gruss JS: Differential diagnosis of the trapezoid-shaped head. Cleft Palate Craniofac J. 2004, 41:13–19. 10.1597/02-053

2. Delashaw JB, Persing JA, Broaddus WC, Jane JA: Cranial vault growth in craniosynostosis. J Neurosurg. 1989, 70:159–165. 10.3171/jns.1989.70.2.0159

3. Johnson D, Wilkie AO: Craniosynostosis. Eur J Hum Genet. 2011, 19:369–376. 10.1038/ejhg.2010.235

4. Jung BK, Yun IS: Diagnosis and treatment of positional plagiocephaly. Arch Craniofac Surg. 2020, 21:80–86. 10.7181/acfs.2020.00059

5. Holowka MA, Reisner A, Giavedoni B, Lombardo JR, Coulter C: Plagiocephaly Severity Scale to Aid in Clinical Treatment Recommendations. J Craniofac Surg. 2017, 28:717–722. 10.1097/SCS.0000000000003520

6. Kalra R, Walker ML: Posterior plagiocephaly. Childs Nerv Syst. 2012, 28:1389–1393. 10.1007/s00381-012-1784-y

7. Marshall JM, Shahzad F: Safe Sleep, Plagiocephaly, and Brachycephaly: Assessment, Risks, Treatment, and When to Refer. Pediatr Ann. 2020, 49:e440–447. 10.3928/19382359-20200922-02

8. Kuo AA, Tritasavit S, Graham JM Jr: Congenital muscular torticollis and positional plagiocephaly. Pediatr Rev. 2014, 35:79–87. 10.1542/pir.35-2-79

9. Van Cruchten C, Feijen MMW, Van Der Hulst R: Demographics of Positional Plagiocephaly and Brachycephaly; Risk Factors and Treatment. J Craniofac Surg 32. 2021, 32:2736–2740. 10.1097/SCS.0000000000007811

10. Van Vlimmeren LA, Van Der Graaf Y, Boere-Boonekamp MM, et al.: Risk factors for deformational plagiocephaly at birth and at 7 weeks of age: a prospective cohort study. Pediatrics 119. 2007, 119:e408–418. 10.1542/peds.2006-2012

11. Joganic JL, Lynch JM, Littlefield TR, Verrelli BC: Risk factors associated with deformational plagiocephaly. Pediatrics. 2009, 124:e1126–1133. 10.1542/peds.2008-2969

12. Oh AK, Hoy EA, Rogers GF: Predictors of severity in deformational plagiocephaly. J Craniofac Surg. 2009, 20:685–689. 10.1097/SCS.0b013e318193d6e5

13. De Luca F, Hinde A: Effectiveness of the ’Back-to-Sleep’ campaigns among healthcare professionals in the past 20 years: a systematic review. BMJ Open. 2016, 6:e011435. 10.1136/bmjopen-2016-011435

14. Turk AE, Mccarthy JG, Thorne CH, Wisoff JH: The “back to sleep campaign” and deformational plagiocephaly: is there cause for concern?. J Craniofac Surg. 1996, 7:12–18. 10.1097/00001665-199601000-00006

15. Pelligra R, Doman G, Leisman G: A reassessment of the SIDS Back to Sleep Campaign. ScientificWorldJournal. 2005, 5:550–557. 10.1100/tsw.2005.71

16. Esparza J, Hinojosa J, Garcia-Recuero I, et al.: Surgical treatment of isolated and syndromic craniosynostosis. Results and complications in 283 consecutive cases. Neurocirugia (Astur). 2008, 19:509-529. 10.1016/s1130-1473(08)70201-x

17. Roby BB, Finkelstein M, Tibesar RJ, Sidman JD: Prevalence of positional plagiocephaly in teens born after the “Back to Sleep” campaign. Otolaryngol Head Neck Surg. 2012, 146:823–828. 10.1177/0194599811434261

18. Collett B, Breiger D, King D, Cunningham M, Speltz M: Neurodevelopmental implications of “deformational” plagiocephaly. J Dev Behav Pediatr. J Dev Behav Pediatr, 26:379–389. 10.1097/00004703-200510000-00008

19. Feijen M, Franssen B, Vincken N, Van Der Hulst RR: Prevalence and Consequences of Positional Plagiocephaly and Brachycephaly. J Craniofac Surg. 2015, 26:e770–773. 10.1097/SCS.0000000000002222

20. Van Wijk RM, Van Vlimmeren LA, Groothuis-Oudshoorn CG, et al.: Helmet therapy in infants with positional skull deformation: randomised controlled trial. BMJ. 2014, 2741:g2741. 10.1136/bmj.g2741

21. Di Chiara A, La Rosa E, Ramieri V, Vellone V, Cascone P: Treatment of Deformational Plagiocephaly With Physiotherapy. J Craniofac Surg. 2019, 30:2008–2013. 10.1097/SCS.0000000000005665

22. Cumpston M, Li T, Page MJ, et al.: Updated guidance for trusted systematic reviews: a new edition of the Cochrane Handbook for Systematic Reviews of Interventions. Cochrane Database Syst Rev. 2019, 10:ED000142. 10.1002/14651858.ED000142

23. Moher D, Liberati A, Tetzlaff J, Altman DG, Group P: Preferred reporting items for systematic reviews and meta-analyses: the PRISMA statement. PLoS Med. 2009, 6:e1000097. 10.1371/journal.pmed.1000097

24. Page MJ, Mckenzie JE, Bossuyt PM, et al.: The PRISMA 2020 statement: an updated guideline for reporting systematic reviews. BMJ. 2021, 372:n71. 10.1136/bmj.n71

25. Bramer WM, Giustini D, Kramer BM: Comparing the coverage, recall, and precision of searches for 120 systematic reviews in Embase, MEDLINE, and Google Scholar: a prospective study. Syst Rev. 2016, 5:39. 10.1186/s13643-016-0215-7

26. Falagas ME, Pitsouni EI, Malietzis GA, Pappas G: Comparison of PubMed, Scopus, Web of Science, and Google Scholar: strengths and weaknesses. FASEB J. 2008, 22:338–342. 10.1096/fj.07-9492LSF

27. Sterne JA, Egger M: Funnel plots for detecting bias in meta-analysis: guidelines on choice of axis. J Clin Epidemiol. 2001, 54:1046–1055. 10.1016/s0895-4356(01)00377-8

28. Aarnivala HE, Valkama AM, Pirttiniemi PM: Cranial shape, size and cervical motion in normal newborns. Early Hum Dev. 2014, 90:425–430. 10.1016/j.earlhumdev.2014.05.007

29. Aarnivala H, Vuollo V, Harila V, et al.: The course of positional cranial deformation from 3 to 12 months of age and associated risk factors: a follow-up with 3D imaging. Eur J Pediatr. 2016, 175:1893–1903. 10.1007/s00431-016-2773-z

30. Ballardini E, Sisti M, Basaglia N, et al.: Prevalence and characteristics of positional plagiocephaly in healthy full-term infants at 8-12 weeks of life. Eur J Pediatr. 2018, 177:1547–1554. 10.1007/s00431-018-3212-0

31. Ifflaender S, Rudiger M, Konstantelos D, Wahls K, Burkhardt W: Prevalence of head deformities in preterm infants at term equivalent age. Early Hum Dev. 2013, 89:1041–1047. 10.1016/j.earlhumdev.2013.08.011

32. Kim EH, Kim KE, Jeon J, et al.: Delayed Motor Development and Infant Obesity as Risk Factors for Severe Deformational Plagiocephaly: A Matched Case-Control Study. Front Pediatr. 2020, 8:582360. 10.3389/fped.2020.582360

33. Leung AYF, Mandrusiak A, Watter P, Gavranich J, Johnston LM: Clinical assessment of head orientation profile development and its relationship with positional plagiocephaly in healthy term infants - A prospective study. Early Hum Dev. 2016, 96:31–38. 10.1016/j.earlhumdev.2016.03.001

34. Maniglio P, Noventa M, Tartaglia S, et al.: The Obstetrician Gynecologist’s role in the screening of infants at risk of severe plagiocephaly: Prevalence and risk factors. Eur J Obstet Gynecol Reprod Biol. 2022, 272:37–42. 10.1016/j.ejogrb.2022.03.011

35. Mawji A, Vollman AR, Fung T, et al.: Risk factors for positional plagiocephaly and appropriate time frames for prevention messaging. Paediatr Child Health. 2014, 19:423–427. 10.1093/pch/19.8.423

36. Nuysink J, Eijsermans MJ, Van Haastert IC, et al.: Clinical course of asymmetric motor performance and deformational plagiocephaly in very preterm infants. J Pediatr. 163, 163:658–665. 10.1016/j.jpeds.2013.04.015

37. Nuysink J, Van Haastert IC, Eijsermans MJ, et al.: Prevalence and predictors of idiopathic asymmetry in infants born preterm. Early Hum Dev. 2012, 88:387–392. 10.1016/j.earlhumdev.2011.10.001

38. Pogliani L, Cerini C, Vivaldo T, Duca P, Zuccotti GV: Deformational plagiocephaly at birth: an observational study on the role of assisted reproductive technologies. J Matern Fetal Neonatal Med. 2014, 27:270–274. 10.3109/14767058.2013.814629

39. Roberts SaG, Symonds JD, Chawla R, et al.: Positional plagiocephaly following ventriculoperitoneal shunting in neonates and infancy-how serious is it?. Childs Nerv Syst. 2017, 33:275–280. 10.1007/s00381-016-3275-z

40. Sheu SU, Ethen MK, Scheuerle AE, Langlois PH: Investigation into an increase in plagiocephaly in Texas from. Arch Pediatr Adolesc Med. 2011, 165:708–713. 10.1001/archpediatrics.2011.42

41. Solani B, Talebian Ardestani M, Boroumand H, et al.: Risk factors associated with positional plagiocephaly in healthy Iranian infants: a case-control study. Iran J Child Neurol. 2022, 16:85–92. 10.22037/ijcn.v16i1.28524

42. Tang M, Gorbutt KA, Peethambaran A, Yang L, Nelson VS, Chang KW: High prevalence of cranial asymmetry exists in infants with neonatal brachial plexus palsy. J Pediatr Rehabil Med. 2016, 9:271–277. 10.3233/PRM-160396

43. Valkama AM, Aarnivala HI, Sato K, et al.: Plagiocephaly after Neonatal Developmental Dysplasia of the Hip at School Age. J Clin Med. 2019, 9:21. 10.3390/jcm9010021

44. Van Vlimmeren LA, Engelbert RH, Pelsma M, et al.: The course of skull deformation from birth to 5 years of age: a prospective cohort study. Eur J Pediatr. 2017, 176:11–21. 10.1007/s00431-016-2800-0

45. Weernink MG, Van Wijk RM, Groothuis-Oudshoorn CG, et al.: Insufficient vitamin D supplement use during pregnancy and early childhood: a risk factor for positional skull deformation. Matern Child Nutr. 2016, 12:177–188. 10.1111/mcn.12153

46. Cavalier A, Picot MC, Artiaga C, et al.: Prevention of deformational plagiocephaly in neonates. Early Hum Dev. 2011, 87:537–543. 10.1016/j.earlhumdev.2011.04.007

47. Pawley N, Bishop NJ: Prenatal and infant predictors of bone health: the influence of vitamin D. Am J Clin Nutr. 2004, 80:1748S–1751S. 10.1093/ajcn/80.6.1748S

48. Vles J, Van Zutphen S, Hasaart T, Dassen W, Lodder J: Supine and prone head orientation preference in term infants. Brain Dev 13. 1991, 13:87–90. 10.1624/105812404X109357

49. Hopkins B, Lems YL, Van Wulfften Palthe T, et al.: Development of head position preference during early infancy: a longitudinal study in the daily life situation. Dev Psychobiol. 1990, 23:39–53. 10.1002/dev.420230105

50. Ronnqvist L, Hopkins B: Head position preference in the human newborn: a new look. Child Dev. 1998, 69:13–23. 10.1111/j.1467-8624.1998.tb06129.x

51. Losee JE, Mason AC, Dudas J, Hua LB, Mooney MP: Nonsynostotic occipital plagiocephaly: factors impacting onset, treatment, and outcomes. Plast Reconstr Surg. 2007, 119:1866–1873. 10.1097/01.prs.0000259190.56177.ca

52. Glasgow TS, Siddiqi F, Hoff C, Young PC: Deformational plagiocephaly: development of an objective measure and determination of its prevalence in primary care. J Craniofac Surg 18. 2007, 18:85–92. 10.1097/01.scs.0000244919.69264.bf

53. Van Vlimmeren LA, Helders PJ, Van Adrichem LN, Engelbert RH: Torticollis and plagiocephaly in infancy: therapeutic strategies. Pediatr Rehabil. 2006, 9:40–46. 10.1080/13638490500037904

54. Greve KR, Goldsbury CM, Simmons EA: Infants With Congenital Muscular Torticollis Requiring Supplemental Physical Therapy Interventions. Pediatr Phys Ther. 2022, 34:335–341. 10.1097/PEP.0000000000000906

55. Ardic C, Usta O, Omar E, Yildiz C, Memis E: Effects of infant feeding practices and maternal characteristics on early childhood obesity. Arch Argent Pediatr. 2019, 117:26–33. 10.5546/aap.2019.eng.26

56. Ross CE, Mirowsky J: The interaction of personal and parental education on health. Soc Sci Med. 2011, 72:591–599. 10.1016/j.socscimed.2010.11.028

57. Moon RY, Locasale-Crouch J, Turnbull KLP, et al.: Investigating Mechanisms for Maternal Education Disparities in Enacting Health-Promoting Infant Care Practices. Acad Pediatr. 2020, 20:926–933. 10.1016/j.acap.2020.03.008

58. Lennartsson F, Nordin P: Nonsynostotic plagiocephaly: a child health care intervention in Skaraborg, Sweden. BMC Pediatr. 2019, 19:48. 10.1186/s12887-019-1405-y

59. Marchac A, Arnaud E, Di Rocco F, Michienzi J, Renier D: Severe deformational plagiocephaly: long-term results of surgical treatment. J Craniofac Surg. 2011, 22:24–29. 10.1097/SCS.0b013e3181f7dd4a

60. Duval S, Tweedie R: Trim and fill: A simple funnel-plot-based method of testing and adjusting for publication bias in meta-analysis. Biometrics. 2000, 56:455–463. 10.1111/j.0006-341x.2000.00455.x

61. Bialocerkowski AE, Vladusic SL, Wei Ng C: Prevalence, risk factors, and natural history of positional plagiocephaly: a systematic review. Dev Med Child Neurol. 2008, 50:577–586. 10.1111/j.1469-8749.2008.03029.x

62. De Bock F, Braun V, Renz-Polster H: Deformational plagiocephaly in normal infants: a systematic review of causes and hypotheses. Arch Dis Child. 2017, 102:535–542. 10.1136/archdischild-2016-312018

63. Inchingolo AI Am, Piras F, Malcangi G, et al.: A Systematic Review of Positional Plagiocephaly Prevention Methods for Patients in Development. Appl Sci. 2022, 12:10.3390/app122111172

64. Martiniuk AL, Vujovich-Dunn C, Park M, Yu W, Lucas BR: Plagiocephaly and Developmental Delay: A Systematic Review. J Dev Behav Pediatr. 2017, 38:67–78. 10.1097/DBP.0000000000000376

65. Rohde JF, Goyal NK, Slovin SR, et al.: Association of Positional Plagiocephaly and Developmental Delay Within a Primary Care Network. J Dev Behav Pediatr. 2021, 42:128–134. 10.1097/DBP.0000000000000860

66. Fontana SC, Daniels D, Greaves T, et al.: Assessment of Deformational Plagiocephaly Severity and Neonatal Developmental Delay. J Craniofac Surg. 2016, 27:1934–1936. 10.1097/SCS.0000000000003014

67. Andrews BT, Fontana SC: Correlative vs. Causative Relationship between Neonatal Cranial Head Shape Anomalies and Early Developmental Delays. Front Neurosci. 2017, 11:708. 10.3389/fnins.2017.00708

